# The Risk Estimation by the PREVENT equations and Clinical Outcome in Patients with Heart Failure with Preserved Ejection Fraction

**DOI:** 10.64898/2026.06.23.26356391

**Authors:** Katsuomi Iwakura, Nobuaki Tanaka, Masato Okada, Akito Nakagawa, Shunsuke Tamaki, Masahiro Seo, Takahisa Yamada, Masamichi Yano, Takaharu Hayashi, Yoshio Yasumura, Yusuke Nakagawa, Katsuki Okada, Yohei Sotomi, Shungo Hikoso, Daisaku Nakatani, Yasushi Sakata, PURUSUIT-HFpEF investigators

**Author notes:** **Corresponding Author:** Katsuomi Iwakura, MD, PhD, Division of Cardiology, Sakurabashi Watanabe Advanced Healthcare Hospital, 4-3-51, Nakanoshima, Kita-ku, Osaka 5300005 Japan.

## Abstract

**Background:** The PREVENT (Predicting Risk of CVD EVENTs) equations estimate the risk of incident cardiovascular disease (CVD) in primary prevention patients. We hypothesized that risk factors incorporated in the equations may be relevant to prognosis in heart failure (HF) and investigated the association between estimated CVD risk and clinical outcomes in patients with preserved ejection fraction (HFpEF).

**Methods:** We estimated the 10-year CVD risk using the PREVENT equations in 278 patients hospitalized for acutely decompensated HFpEF (median 75 years, 51.4% male). We divided them into four groups according to the quartiles of estimated CVD risk and followed them to observe major adverse cardiovascular events (MACE), a composite of all-cause death, HF hospitalization, and stroke.

**Results:** MACE occurred in 125 patients (45.0%) over a median follow-up of 1,050 days. The estimated CVD risk classification was an independent predictor for MACE (p=0.02) in the multivariable Cox proportional hazard model. There was a difference in MACE-free survival across the four quartile groups (p<0.001 by log-rank test), and the lowest CVD risk group had significantly lower MACE incidence than other groups. The estimated CVD risk provided incremental prognostic value beyond N-terminal pro-B type natriuretic peptide (C-index: 0.626 vs, P=0.009). The predictive value of the estimated CVD risk for MACE at 1 year was comparable to that of the MAGGIC score (AUC 0.676 vs. 0.639, p=0.42).

**Conclusions:** The 10-year CVD risk estimated by the PREVENT equations had a moderate predictive value for MACE in patients hospitalized for HFpEF.

---

Prevalence of heart failure (HF) is globally rising, and HF with preserved ejection fraction (HFpEF) is becoming the dominant subtype^1^. The prevalence of HFpEF relative to HF with reduced ejection fraction (HFrEF) is increasing at a rate of 1% per year, and HFpEF now accounts for approximately half of all HF cases^1,2^. The rising prevalence of HFpEF is attributed to multiple factors including population aging, increasing obesity rates, rising diabetes prevalence, and improved survival from myocardial infarction and other comorbidities^2–4^. While the prognosis of HF has improved significantly through advances in therapy and management, improvement in HFpEF prognosis have lagged behind those of HFrEF^1,3^. Patients with HFpEF are hospitalized approximately 1.4 times per year and have an annual mortality rate of approximately 15%^4^. Routine assessment of risk for adverse outcomes in HF patients was recommended by guidelines^5^, and general HF risk scores such as the Barcelona Bio-Heart Failure (BCN-Bio-HF) risk calculator^6^, the MAGGIC (Meta-Analysis Global Group in Chronic Heart Failure) score^7,8^ and Seattle Heart Failure Model (SHFM)^9^ can be applied to patients with HFpEF.

American Heart Association developed the PREVENT (Predicting Risk of Cardiovascular Disease Events) equations to estimate 10-year and 30-year risk for cardiovascular disease (CVD) in adults aged 30 to 79 years^10,11^. They are an update of the 2013 pooled cohort equations (PCEs) aimed at achieving more accurate CVD risk estimation in contemporary populations. Compared with the PCEs, the PREVENT equations expand the age range to 30–79 years, add HF to the predicted outcomes, add kidney and metabolic factors, and remove race from the calculation. The PREVENT equations provide separate risk estimates for three outcomes: atherosclerotic cardiovascular disease (ASCVD) alone, HF alone, and total CVD. They are calculated using routinely available clinical variables such as age, sex, blood pressure, total and high-density lipoprotein (HDL) cholesterol, diabetes status, smoking, estimated glomerular filtration rate (eGFR), statin use, and antihypertensive medication use. They demonstrate excellent discrimination and significantly improved calibration compared to the PCEs^10,11^. The PREVENT equations are based on race-free cardiovascular risk prediction models, and have been shown to predict total CVD, ASCVD, and HF risk in Asian population as well as in the original evaluation studies^12,13^.

While the PREVENT equations are designed for risk estimation and primary prevention in individuals without a history of CVD, they incorporate the clinical factors which may be relevant to the prognosis of patients with established HF. In this study, we retrospectively investigated the association between estimated CVD risk and clinical outcomes in patients hospitalized for acutely decompensated HFpEF.

## Methods

### Study Population

We retrospectively examined patients registered in the Prospective Multicenter Observational Study of Patients with Heart Failure with Preserved Ejection Fraction (PURSUIT-HFpEF), and patients aged 30 to 79 years were enrolled for the present study. The PURSUIT-HFpEF study is a multicenter, observational study enrolling consecutive patients hospitalized with acute decompensated HFpEF (left ventricular ejection fraction (LVEF) ≥ 50%) in 31 collaborating hospitals [UMIN-CTR ID: UMIN000021831]. Details of the entry criteria and data collection have been described elsewhere^14^. Briefly, patients admitted with acutely decompensated HFpEF were registered, and their clinical data including medications, laboratory tests, and ECG were collected on admission, at discharge, and 1 year after discharge. Acutely decompensated HFpEF was diagnosed based on the following criteria: (1) clinical symptoms and signs according to the Framingham Heart Study criteria, (2) LVEF on admission ≥50%, and (3) serum N-terminal pro-B type natriuretic peptide (NT-proBNP) ≥400 pg/mL or brain natriuretic peptide ≥100 pg/mL^14^.

We excluded patients whose variables were missing or out of range for risk calculations; total cholesterol must be within the range of 30 to 320 mg/dL, HDL cholesterol 20 to 100 mg/dL, systolic blood pressure 90 to 200 mm Hg, and body mass index (BMI) 18.5 to less than 40.0^11^. Those who died during the index hospitalization were also excluded.

Study patients were followed by direct contact or telephone interview to observe major adverse cardiovascular events (MACE), a composite of all-cause death, heart failure hospitalization, and stroke. For patients whose survival status could not be determined by these means, data from the National Vital Statistics of Japan, which includes all death records in Japan reported by prefectural public health centers, were used with the permission of the Ministry of Health, Labor and Welfare in accordance with the Statistics Act in Japan.

This study was conducted in accordance with the Declaration of Helsinki. The study protocol was approved by the Osaka University Hospital Clinical Research Review Committee and by each corresponding hospital’s Ethics Committee according to the Ethical Guidelines for Medical and Health Research Involving Human Subjects issued by the Ministry of Health, Labor and Welfare in Japan. Informed consent was obtained from each patient by one of the investigators before enrollment.

### Risk Calculation using the PREVENT Equations

Variables used for the PREVENT equations were collected just before the hospital discharge. If multiple measures were available for a parameter, the measurement closest to discharge was used for the calculation. The PREVENT equations estimate the absolute 10-year risk of total CVD as a composite ASCVD and HF, ASCVD alone, and HF alone^11^. We primarily used the 10-year risk of total CVD to predict MACE in the present study. We calculated it using the American Heart Association PREVENT Online Calculator (https://professional.heart.org/en/guidelines-and-statements/prevent-calculator). We also calculated the 10-year ASCVD and HF risk to compare the predictive value of these estimated risks.

The base PREVENT model includes age, sex, smoking status, systolic blood pressure, total cholesterol, HDL cholesterol, diabetes status, antihypertensive medication use, statin use, and eGFR. Hemoglobin A1c (Hb A1c), urine albumin-to-creatinine ratio, and the social deprivation index are optional for further personalization of risk assessment when available. In the present study, the risk was calculated using the basic variables and HbA1c values; those whose HbA1c value was missing were not excluded and the risk was calculated without it. Study patients were divided into four groups based on the quartile values of the 10-year CVD risk (Q1 as the lowest and Q4 as the highest risk).

### Calculation of the MAGGIC Score

To compare the predictive ability of the PREVNT 10-year CVD risk, the MAGGIC risk score was calculated for those with available data using the following variables; left ventricular ejection fraction (LVEF), age, systolic blood pressure, BMI, creatinine, New York Heart Association (NYHA) class, male sex, current smoking, diabetes, a diagnosis of chronic obstructive pulmonary disease, a first diagnosis of heart failure in the 18 months, and the absence of β-blockers and not on renin-angiotensin-aldosterone system (RAS) inhibitors use ^8^.

### Statistical Analysis

The normality of the distribution of continuous variables was assessed using the Kolmogorov-Smirnov test. Normally distributed variables are presented as the mean ± standard deviation and non-normally distributed variables as the median (interquartile range). Comparisons were made using the Student’s T test or one-way ANOVA for normally distributed variables, and the Mann-Whitney U test or the Kruskal-Wallis test for non-normally distributed variables. Categorical variables were compared with Fisher’s exact test, and p-values were determined using a simulation approach because of the large amount of data. Pairwise comparisons were performed using the Bonferroni correction when statistical significance was observed among the groups.

The quartile classification of CVD risk was included as categorical variables in the univariable and multivariable Cox proportional hazards models. Univariable Cox proportional hazard models for MACE, all-cause death, and HF hospitalization were constructed using the quartiles of CVD score, and the relevant variables such as age, gender, BMI, history of diabetes, hypertension, dyslipidemia, use of diuretics, RAS inhibitors, mineral corticoid receptor antagonists (MRA), β-blockers, and statins, and laboratory values at discharge. N-terminal pro-B-type natriuretic peptide (NT-proBNP) was log-transformed (LN(NT-proBNP)) for the analysis. Multivariable Cox regression models were constructed using variables showing P<0.05 in the univariable analysis. Event-free survival analysis was performed using the Kaplan-Meier method with the log-rank test for comparisons among the four groups based on the quartiles of estimated CVD risk.

To evaluate the incremental prognostic value of the PREVNT 10-year CVD risk for MACE, we constructed two nested Cox proportional hazards models: Model 1 included (LN(NT-proBNP)) alone, and Model 2 additionally incorporated 10-year CVD risk. The incremental prognostic value of 10-year CVD risk was evaluated by comparing model discrimination using Harrell’s concordance index (C-index) and differences in C-index between two models were assessed using the compareC method. Model fit was additionally evaluated using likelihood ratio testing and Akaike information criterion (AIC).To compare the predictive values of the CVD risk and the MAGGIC scores, or between the CVD risk and the ASCVD or HF risk, time-dependent receiver operating characteristic (ROC) curves for clinical outcomes at 1 year were constructed and the area under the curves (AUCs) were compared. All statistical analyses were performed using R (R Foundation for Statistical Computing, Vienna, Austria) and R with a graphical user interface EZR (Saitama Medical Centre, Jichi Medical University, Japan).

## Results

### Patient Characteristics

Among 1,238 consecutive patients (median age 83 (77, 87) years) registered in the PURSUIT-HFpEF study between June 2016 and February 2022, 433 patients aged 30 to 79 years were initially enrolled for the present study. All patients excluded due to age were >79 years. Of these, 153 (35.3%) patients were excluded due to in-hospital death (4 patients), missing variables needed for the CVD risk calculation (13 patients), no follow-up after discharge (10 patients), and variables outside the acceptable range for CVD risk calculation (128 patients): total cholesterol in 42 patients (all <130mg/dL), systolic blood pressure in 7 patients, BMI in 58 patients (all <18.5kg/m^2^), and eGFR in 36 patients. Some patients had more than one variable outside the acceptable range for calculation, and the total number of patients excluded for out-of-range variables was smaller than the sum of exclusions reported for each variable (Figure 1). We calculated the 10-year CVD risk in the remaining 278 patients (age 75 (71, 77) years, 51.4% male (n=143), BMI 25.3 (22.7, 29.8) kg/m^2^). HbA1c was included in the calculation in 228 patients (82.0%). Patients excluded from the analysis had significantly lower BMI, hematologic indices, albumin, renal function, and lipid levels, and higher NT-proBNP levels, whereas age, sex, and the prevalence of comorbidities, and most medications did not differ significantly (Supplement Table S1).

**Figure 1.**
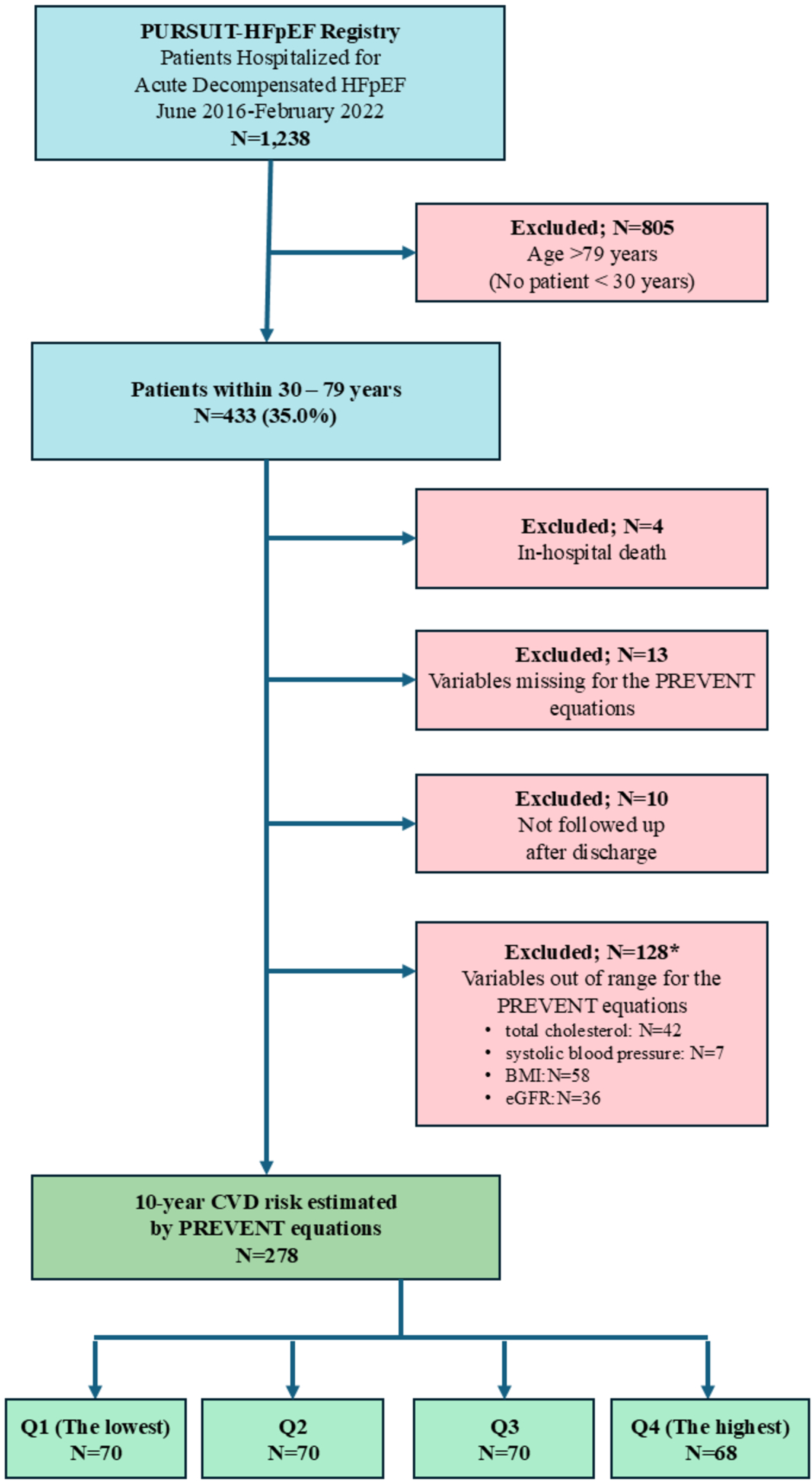
CONSORT Flow Diagram of Study Population Selection. From the PURSUIT-HFpEF Registry of 1,238 patients hospitalized for acute decompensated HFpEF, 433 patients aged 30 to 79 years were initially enrolled for the present study. A total of 153 (35.3%) patients were excluded for the following reasons; in-hospital death (4 patients), missing variables for the PREVENT equations (13 patients), no follow-up data (10 patients), and variables outside the acceptable range for PREVENT risk calculation (128 patients; total cholesterol, 42 patients; systolic blood pressure, 7 patients; BMI, 58 patients; eGFR, 36 patients). Some patients had more than one out-of-range variable, and the category counts do not sum to the total number of excluded patients. The 10-year total CVD risk was estimated in the remaining 278 patients, who were divided into four groups according to the quartiles of estimated CVD risk. HFpEF denotes heart failure with preserved ejection fraction; BMI, body mass index; eGFR, estimated glomerular filtration rate; CVD, cardiovascular disease; PREVENT, Predicting Risk of Cardiovascular Disease Events.

The mean 10-year CVD risk in the study patients was 29.1±9.6 %, and they were divided into four groups based on the quartiles of CVD risk (Q1, n=70, range 0.8 - 22.6 %; Q2. n=70, 22.7 - 29.6 %; Q3, n=70, 29.7 - 35.1 %: Q4, n=68, 35.2 - 57.5 %). There were several differences in clinical background and laboratory findings between the four groups. Patients with higher CVD risk were older and had higher BMI and higher prevalence of comorbidities such as hypertension, diabetes, and dyslipidemia (Table 1). Diuretics were more frequently administered to patients with higher CVD risk (p=0.01). No significant difference was observed in other medications such as RAS inhibitors, MRA, and β-blockers, except for statins. Patients with higher CVD risk had lower kidney function, and higher NT-proBNP. In addition, there were significant differences in triglyceride, blood glucose, and HbA1c between the four groups (Table 1).

**Table 1.** Baseline characteristics of patients stratified by estimated 10-year CVD risk.

| Variable | All patients | Classification by the quartile of 10-year CVD risk |  |  |  | p |
| --- | --- | --- | --- | --- | --- | --- |
|  |  | Q1 | Q2 | Q3 | Q4 |  |
| **Number** | 278 | 70 | 70 | 70 | 68 |  |
| **10-year risk of CVD, %**<br>**(range)** | 29.0 ± 9.6 | 17.1 ± 5.5*<sup>‡\|\|</sup><br>(0.8 - 22.6) | 25.9 ± 2.1*<sup>‡</sup><br>(22.7 - 29.6) | 32.3 ± 1.6*<br>(29.7 - 35.1) | 41.1 ± 5.1<br>(35.2 - 57.5) | <0.001 |
| **Age, year** | 75 (71-77) | 70 (64-73)*<sup>‡\|\|</sup> | 75 (72-76)<sup>†§</sup> | 76 (73-78) | 77 (73-78) | <0.001 |
| **Male, n (%)** | 143 (51.4) | 32 (45.7) | 28 (40)<sup>†</sup> | 38 (54.3) | 45 (66.2) | 0.01 |
| **Body Mass index, kg/m<sup>2</sup>** | 25.3 (22.7-29.8) | 24.8 (22.3-28.9)<sup>†</sup> | 24.1 (22.3-28.5)* | 25.6 (23.0-27.8) | 27.3 (23.8-31.1) | 0.002 |
| **Systolic BP, mmHg** | 122 ± 17 | 120 ± 15 | 119 ± 16 | 122 ± 17 | 125 ± 20 | 0.22 |
| **Hypertension, n (%)** | 233 (83.8) | 49 (70.0)*<sup>‡</sup> | 57 (81.4) | 64 (92.8) | 63 (92.8) | <0.001 |
| **Diabetes, n (%)** | 120 (43.2) | 15 (21.4)*<sup>‡</sup> | 22 (31.4)* | 36 (51.4) | 47 (69.1) | <0.001 |
| **Dyslipidemia, n (%)** | 127 (45.7) | 30 (43.5) | 27 (39.1)<sup>†</sup> | 28 (40.0) | 42 (62.7) | 0.02 |
| **Current smoker, n (%)** | 52 (18.7) | 12 (17.1) | 8 (11.4)<sup>†</sup> | 10 (14.3) | 22 (32.4) | 0.01 |
| **Antihypertensive drugs, n (%)** | 270 (97.1) | 63 (90) | 69 (98.6) | 70 (100) | 68 (100) | 0.001 |
| **RAS inhibitors, n (%)** | 162 (58.3) | 38 (54.3) | 38 (54.3) | 44 (62.9) | 42 (61.8) | 0.61 |
| **β-blockers, n (%)** | 166 (59.7) | 38 (54.3) | 44 (62.9) | 42 (60.0) | 42(61.8) | 0.75 |
| **MRA, n (%)** | 113 (40.7) | 29 (41.4) | 35 (50.0) | 28 (40.0) | 21 (30.9) | 0.16 |
| **Statins, n (%)** | 91 (32.7) | 21 (30.0) | 26 (37;1) | 20 (28.6) | 24 (35.3) | 0.66 |
| **Diuretics, n (%)** | 215 (77.3) | 45 (64.3)<sup>†</sup> | 53 (75.7) | 58 (82.9) | 59 (86.8) | 0.01 |
| **Red blood cell, 10<sup>6</sup>/μl** | 4.11 ± 0.67 | 4.08 ± 0.74 | 4.13 ± 0.62 | 4.23 ± 0.67 | 4.01 ± 0.64 | 0.26 |
| **Hemoglobin, g/dl** | 12.3 ± 2.1 | 12.5 ± 2.1 | 12.5 ± 2.0 | 12.3 ± 2.2 | 12.0 ± 2.1 | 0.58 |
| **Hematocrit, %** | 37.3 ± 6.1 | 37.4 ± 6.4 | 37.8 ± 5.9 | 37.7 ± 6.0 | 36.4 ± 6.0 | 0.53 |
| **White blood cell, 10<sup>3</sup>/μl** | 5.90 (4.70-7.00) | 5.80 (4.53-6.70) | 5.95 (4.70-6.97) | 5.70 (4.55-7.15) | 6.10 (5.10-7.30) | 0.25 |
| **Platelets, 10<sup>4</sup>/μl** | 22.5 (17.1-29.2) | 22.3 (18.0-28.3) | 21.3 (16.1-29.0) | 24.1 (17.1-29.3) | 22.5 (17.9-29.2) | 0.41 |
| **Total protein, g/dl** | 6.8 ± 0.7 | 6.8 ± 0.7 | 6.8 ± 0.7 | 6.7 ± 0.6 | 6.8 ± 0.8 | 0.93 |
| **Albumin, g/dl** | 3.6 (3.3-3.8) | 3.5 (3.3-3.8) | 3.5 (3.3-3.8) | 3.6 (3.3-3.8) | 3.6 (3.3-3.8) | 0.99 |
| **AST, U/l** | 22.0 (17.0-30.0) | 21.5 (17.0-28.8) | 25.5 (18.3-32.0)<sup>†</sup> | 23.0 (17.5-31.0) | 20.0 (15.0-26.0) | 0.03 |
| **ALT, U/l** | 16.0 (11.0-26.0) | 16.0 (12.0-27.8) | 18.5 (13.3-28.0) | 16.0 (12.0-25.5) | 14.0 (10.0-21.0) | 0.06 |
| **Sodium, mEq/L** | 140 (138-141) | 140 (139-141) | 140 (138-142) | 140 (138-142) | 139 (138-141) | 0.38 |
| **Potassium, mEq/L** | 4.3 (3.9-4.6) | 4.2 (3.9-4.4) | 4.3 (3.9-4.6) | 4.2 (3.9-4.6) | 4.40 (3.8-4.7) | 0.19 |
| **Chloride, mEq/L** | 104 (101-106) | 104 (102-106) | 103 (99-106) | 104 (101-106) | 103 (100-106) | 0.28 |
| **Blood urea, mg/dl** | 21;9 (16.9-28.9) | 17.4 (15.0-22.8)*<sup>§</sup> | 21.0 (16.1-26.9)* | 20.7 (17.2-28.5)* | 28.2 (22.8-36.2) | <0.001 |
| **Creatinine, mg/dl** | 1.0 (0.8-1.3) | 0.9 (0.7-1.0)*<sup>‡</sup> | 1.0 (0.7-1.1)*<sup>§</sup> | 1.1 (0.9-1.3)* | 1.6 (1.3-2.1) | <0.001 |
| **eGFR, mL/min/1.73m<sup>2</sup>** | 46.6 (37.0-61.1) | 60.3 (48.5-72.9)*<sup>‡</sup> | 55.4 (41.7-62.3)* | 46.2 (37.9-54.9)* | 32.5 (23.0-41.4) | <0.001 |
| **Uric acid, mg/dl** | 7.1 ± 1.9 | 6.7 ± 1.7<sup>†</sup> | 6.7 ± 1.8<sup>†</sup> | 7.1 ± 2.1 | 7.8 ± 1.8 | <0.001 |
| **Total cholesterol, mg/dl** | 166 (148-186) | 173 (154-197) | 164 (148-185) | 165 (145-185) | 163 (148-180) | 0.2 |
| **Triglyceride, mg/dl** | 111 (86-146) | 112 (82.5-142)<sup>†</sup> | 100 (75.8-119.3)* | 108 (80-141.5)<sup>†</sup> | 136 (96-183.5) | <0.001 |
| **HDL-C, mg/dl** | 42 (36-50) | 43.5 (37.0-51.0) | 45.0 (38.0-55.0)* | 42 (35-47) | 39 (32-48) | 0.005 |
| **LDL-C, mg/dl** | 100 (85-118) | 107.0 (87.3-122.8) | 101 (80-113) | 100.0 (86.3-120.8) | 97.0 (83.8-111.0) | 0.16 |
| **Blood sugar, mg/dl** | 99 (89-119) | 95 (87-108)* | 97 (89-105)* | 97 (87-112)* | 113 (96-142) | <0.001 |
| **HbA1c, %** | 6.1 (5.7-6.6) | 5.7 (5.5-6.0)*§ | 5.9 (5.6-6.4)* | 6.1 (5.7-6.6)* | 6.8 (6.2-7.5) | <0.001 |
| **C-reactive protein, mg/l** | 2.7 (1.0-7.6) | 2.6 (0.9-7.9) | 3.0 (1.2-8.2) | 3.3 (1.0-6.5) | 2.2 (0.9-5.6) | 0.83 |
| **NT-proBNP, pg/ml** | 724.5<br>(341.7-1648.5) | 575.0<br>(195.5-1270.0)* | 735.0<br>(406.0-1578.0) | 515.5<br>(334.3-1705.3) | 925.0<br>(537.3-2263.2) | 0.007 |
| **LN(NT-proBNP)** | 6.58 ± 1.12 | 6.17 ± 1.16* | 6.58 ± 1.02 | 6.58 ± 1.16 | 6.94 ± 1.04 | 0.002 |

CVD denotes cardiovascular disease; BP, blood pressure; RAS, renin angiotensin system; MRA, mineral corticoid receptor antagonists; AST, aspartate aminotransferase; ALT, alanine aminotransferase; eGFR, estimated glomerular filtration rate; HDL-c, high-density lipoprotein cholesterol; LDL-c, low-density lipoprotein cholesterol; HbA1c, hemoglobin A1c; NT-proBNP, N-terminal pro-brain natriuretic peptide; LN(NT-proBNP), logarithm of N-terminal pro-brain natriuretic peptide.
\* p<0.01, † p<0.05 vs. Q4; ‡p<0.01, §: p<0.05 vs. Q3; ||:p<0.01 vs. Q2

### Association between CVD Risk and MACE

During a follow up period of a median of 1050 days (IQR 434 - 1468 days), MACE was observed in 125 patients (45.0%). All-cause death was observed in 58 patients (20.9%), HF hospitalization in 83 patients (29.9%), and stroke in 21 patients (7.6%). Univariable Cox proportional hazard analysis identified diabetes, dyslipidemia, β-blockers, diuretics, serum albumin, blood urea nitrogen (BUN), serum creatinine, eGFR, LN(NT-proBNP), and the CVD risk classification as significant predictors for MACE, and they were included into the multivariable Cox model (Table 2). Red blood cell count, hemoglobin, and hematocrit were also selected as independent predictors, and due to their strong intercorrelation, hemoglobin was included in a multivariable analysis as a representative variable. Among these variables, serum creatinine, LN(NT-proBNP), eGFR, and the CVD risk classification were identified as independent predictors of MACE (p=0.02). When Q1 of the CVD risk was used as the reference, Q2 and Q3 had significantly higher incidence of MACE (Q2: HR 2.83, 95% CI 1.39 – 5.74, p=0.004; Q3: HR 2.52, 95% CI 1.25-5.06, p=0.01), while the difference between Q1 and Q4 did not reach the statistical significance (HR 1.72, 95% CI 0.79 – 3.72, p=0.17). (Table 2). Kaplan-Meier curves demonstrated a significant difference in the incidence of MACE between the four groups (p< 0.001 by log-rank test); and the-lowest risk group (Q1) had a significantly lower incidence of MACE than the remaining three groups (Q2, p=0.004; Q3, p=0.004; Q4, p<0.001 vs. Q1) (Figure 2).

**Figure 2.**
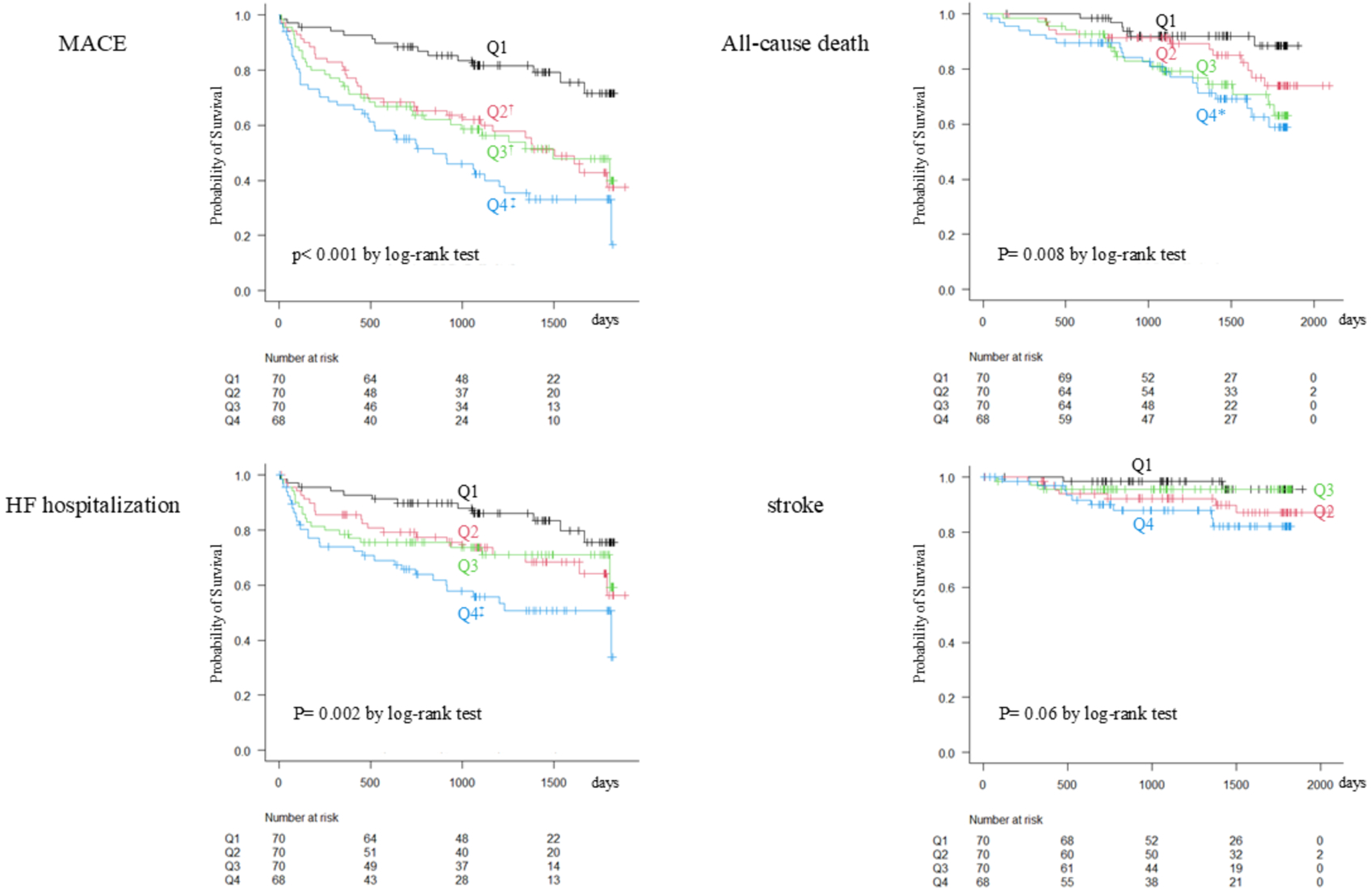
Estimated 10-year CVD risk and Clinical outcomes. Kaplan-Meier curves for MACE, a composite of all-cause death, heart failure (HF) hospitalization, and stroke (upper left), all-cause death (upper right), HF hospitalization (lower left), and stroke (lower right), stratified according to the quartiles of 10-year CVD risk estimated by the PREVENT equation. There were significant differences in the incidence of MACE (p< 0.001 by log-rank test), all-cause death (p= 0.008) and HF hospitalization (p= 0.002) among the four groups, whereas no significant difference was observed in the incidence of stroke (p=0.06). The highest risk group (Q4) had higher incidence of MACE than other three groups, and a higher incidence of both all-cause mortality and HF hospitalization rate than the lowest risk group (Q1). MACE denotes major adverse cardiovascular events: PREVENT, Predicting Risk of Cardiovascular Disease Events. *: p<0.01, †: p<0.05,‡: p<0.001 vs. Q4

**Table 2.** Univariable and multivariable Cox proportional hazards models for MACE.

|  | Univariable |  |  | Multivariable |  |  |
| --- | --- | --- | --- | --- | --- | --- |
|  | HR | 95% CI | p | HR | 95% CI | p |
| **Age** | 1.03 | 1.00-1.05 | 0.051 |  |  |  |
| **Male sex** | 0.81 | 0.57-1.15 | 0.24 |  |  |  |
| **Body Mass Index** | 0.99 | 0.96-1.03 | 0.78 |  |  |  |
| **Systolic Blood pressure** | 1.00 | 0.99-1.01 | 0.63 |  |  |  |
| **Hypertension** | 1.46 | 0.85-2.50 | 0.17 |  |  |  |
| **Dyslipidemia** | 1.50 | 1.05-2.14 | 0.02 | 1.41 | 0.91-2.16 | 0.12 |
| **Diabetes** | 1.56 | 1.10-2.21 | 0.01 | 1.46 | 0.96-2.23 | 0.08 |
| **Current smoker** | 0.84 | 0.52-1.35 | 0.46 |  |  |  |
| **Antihypertensive drugs** | 2.33 | 0.58-9.44 | 0.23 |  |  |  |
| **RAS inhibitors** | 1.11 | 0.77-1.59 | 0.58 |  |  |  |
| **β blockers** | 1.23 | 0.86-1.77 | 0.26 |  |  |  |
| **MRA** | 0.78 | 0.54-1.12 | 0.18 |  |  |  |
| **Statin** | 1.23 | 0.85-1.76 | 0.27 |  |  |  |
| **Diuretics** | 1.63 | 1.01-2.63 | 0.05 | 0.83 | 0.48-1.44 | 0.51 |
| **Red blood cells** | 1.00 | 0.99-1.00 | <0.001 | - | - | - |
| **Hemoglobin** | 0.85 | 0.78-0.93 | <0.001 | 0.91 | 0.82-1.01 | 0.08 |
| **Hematocrit** | 0.95 | 0.92-0.98 | <0.001 | - | - | - |
| **White blood cells** | 1.02 | 0.94-1.11 | 0.63 |  |  |  |
| **Platelets** | 1.01 | 0.99-1.03 | 0.54 |  |  |  |
| **Total protein** | 0.79 | 0.60-1.03 | 0.09 |  |  |  |
| **Albumin** | 0.60 | 0.40-0.90 | 0.01 | 0.79 | 0.47-1.34 | 0.39 |
| **AST** | 1.00 | 0.99-1.02 | 0.54 |  |  |  |
| **ALT** | 1.00 | 0.99-1.01 | 0.40 |  |  |  |
| **Sodium** | 0.98 | 0.92-1.05 | 0.55 |  |  |  |
| **Potassium** | 0.96 | 0.66-1.39 | 0.82 |  |  |  |
| **Chloride** | 0.98 | 0.94-1.03 | 0.49 |  |  |  |
| **Blood urea nitrogen** | 1.03 | 1.02-1.05 | <0.001 | 1.02 | 1.00-1.04 | 0.02 |
| **Creatinine** | 2.45 | 1.81-3.31 | <0.001 | 2.08 | 1.11-3.91 | 0.02 |
| **eGFR** | 0.98 | 0.97-0.99 | <0.001 | 1.02 | 1.00-1.04 | 0.08 |
| **Blood glucose** | 1.00 | 1.00-1.01 | 0.88 |  |  |  |
| **HbA1c** | 1.09 | 0.90-1.33 | 0.37 |  |  |  |
| **Total cholesterol** | 1.00 | 0.99-1.00 | 0.47 |  |  |  |
| **Triglyceride** | 1.00 | 1.00-1.01 | 0.052 |  |  |  |
| **HDL-C** | 1.00 | 0.98-1.01 | 0.73 |  |  |  |
| **LDL-C** | 1.00 | 0.99-1.00 | 0.27 |  |  |  |
| **C-reactive protein** | 1.13 | 0.99-1.29 | 0.06 |  |  |  |
| **LN(NTproBNP)** | 1.58 | 1.33-1.87 | <0.001 | 1.36 | 1.12-1.67 | 0.002 |
| **Quartile of CVD Risk** |  |  | <0.001 |  |  | 0.02 |
| ***Q2*** | 2.68 | 1.46-4.90 | 0.001 | 2.83 | 1.39-5.75 | 0.004 |
| ***Q3*** | 2.71 | 1.47-4.99 | 0.001 | 2.52 | 1.25-5.06 | 0.010 |
| ***Q4*** | 4.10 | 2.27-7.39 | <0.001 | 1.72 | 0.79-3.72 | 0.17 |

MACE denotes major adverse cardiovascular events; RAS, renin angiotensin system; MRA, mineral corticoid receptor antagonists; AST, aspartate aminotransferase; ALT, alanine aminotransferase; eGFR, estimated glomerular filtration rate; HbA1c, hemoglobin A1c; HDL-c, high-density lipoprotein cholesterol; LDL-c, low-density lipoprotein cholesterol; LN(NT-proBNP), logarithm of N-terminal pro-brain natriuretic peptide; CVD, cardiovascular disease.

There were a significant differences in the incidence of all-cause death (p= 0.008 by log-rank test) and that of HF hospitalization (p= 0.002) between the four groups; and the Q1 group had lower incidence than the Q4 group for both outcomes (p=0.008 for all-cause death and p<0.001 for HF hospitalization) (Figure 2). The CVD risk classification was identified as an independent predictor of all-cause death and HF hospitalization in the univariable analysis; however, it was not retained as an independent predictor in the multivariable analysis for either outcome (Table 3 and 4). The difference in the incidence of stroke among the four groups was not significant (p=0.06 by log-rank test), and the CVD risk classification was not identified as an independent predictor in the univariable analysis (Supplementary Table S2).

**Table 3.** Univariable and multivariable Cox proportional hazards models for all-cause death.

|  | Univariable |  |  | Multivariable |  |  |
| --- | --- | --- | --- | --- | --- | --- |
|  | HR | 95% CI | p | HR | 95% CI | p |
| **Age** | 1.08 | 1.02-1.14 | 0.006 | 1.07 | 1.01-1.14 | 0.03 |
| **Male Sex** | 0.72 | 0.43-1.22 | 0.22 |  |  |  |
| **Body Mass Index** | 0.97 | 0.91-1.02 | 0.22 |  |  |  |
| **Systolic Blood pressure** | 1.01 | 0.99-1.02 | 0.32 |  |  |  |
| **Hypertension** | 2.32 | 0.84-6.41 | 0.10 |  |  |  |
| **Dyslipidemia** | 1.51 | 0.89-2.57 | 0.13 |  |  |  |
| **Diabetes** | 1.63 | 0.97-2.74 | 0.06 |  |  |  |
| **Current smoker** | 0.84 | 0.41-1.72 | 0.64 |  |  |  |
| **Antihypertensive drugs** | 1.83 | 0.25-13.19 | 0.55 |  |  |  |
| **RAS inhibitors** | 0.73 | 0.43-1.22 | 0.23 |  |  |  |
| **β blockers** | 0.77 | 0.46-1.29 | 0.32 |  |  |  |
| **MRA** | 0.69 | 0.39-1.20 | 0.19 |  |  |  |
| **Statin** | 0.73 | 0.41-1.28 | 0.27 |  |  |  |
| **Diuretics** | 2.24 | 1.02-4.94 | 0.045 | 1.34 | 0.59-3.08 | 0.49 |
| **Red blood cells** | 1.00 | 0.99-1.00 | 0.01 | - | - | - |
| **Hemoglobin** | 0.84 | 0.74-0.95 | 0.007 | 0.92 | 0.79-1.08 | 0.31 |
| **Hematocrit** | 0.95 | 0.91-0.99 | 0.02 | - | - | - |
| **White blood cells** | 0.96 | 0.83-1.10 | 0.54 |  |  |  |
| **Platelets** | 0.99 | 0.96-1.02 | 0.40 |  |  |  |
| **Total protein** | 0.79 | 0.54-1.17 | 0.24 |  |  |  |
| **Albumin** | 0.52 | 0.30-0.92 | 0.02 | 0.47 | 0.22-1.01 | 0.054 |
| **AST** | 1.01 | 0.99-1.03 | 0.37 |  |  |  |
| **ALT** | 1.00 | 0.99-1.01 | 0.92 |  |  |  |
| **Sodium** | 0.97 | 0.88-1.06 | 0.47 |  |  |  |
| **Potassium** | 0.63 | 0.37-1.09 | 0.10 |  |  |  |
| **Chloride** | 0.95 | 0.88-1.01 | 0.12 |  |  |  |
| **Blood urea nitrogen** | 1.02 | 1.00-1.04 | 0.04 | 1.00 | 0.97-1.04 | 0.84 |
| **Creatinine** | 1.60 | 1.08-2.37 | 0.02 | 1.28 | 0.66-2.49 | 0.46 |
| **eGFR** | 0.99 | 0.98-1.01 | 0.23 |  |  |  |
| **Blood glucose** | 1.00 | 0.99-1.01 | 0.93 |  |  |  |
| **HbA1c** | 1.05 | 0.79-1.40 | 0.72 |  |  |  |
| **Total cholesterol** | 1.00 | 0.99-1.01 | 0.50 |  |  |  |
| **Tryglyceride** | 1.00 | 1.00-1.01 | 0.19 |  |  |  |
| **HDL-C** | 1.02 | 0.99-1.04 | 0.21 |  |  |  |
| **LDL-C** | 1.00 | 0.99-1.01 | 0.84 |  |  |  |
| **C-reactive protein** | 1.11 | 0.92-1.34 | 0.28 |  |  |  |
| **LN(NTproBNP)** | 1.40 | 1.09-1.79 | 0.009 | 1.08 | 0.82-1.43 | 0.58 |
| **Quartile of CVD Risk** |  |  | 0.01 |  |  | 0.93 |
| ***Q2*** | 2.10 | 0.80-5.51 | 0.13 | 1.13 | 0.40-3.20 | 0.82 |
| ***Q3*** | 3.41 | 1.35-8.59 | 0.009 | 1.38 | 0.49-3.87 | 0.54 |
| ***Q4*** | 3.98 | 1.60-9.85 | 0.003 | 1.18 | 0.43-3.25 | 0.74 |

MACE denotes major adverse cardiovascular events; RAS, renin angiotensin system; MRA, mineral corticoid receptor antagonists; AST, aspartate aminotransferase; ALT, alanine aminotransferase; eGFR, estimated glomerular filtration rate; HbA1c, hemoglobin A1c; HDL-c, high-density lipoprotein cholesterol; LDL-c, low-density lipoprotein cholesterol; LN(NT-proBNP), logarithm of N-terminal pro-brain natriuretic peptide; CVD, cardiovascular disease.

**Table 4.** Univariable and multivariable Cox proportional hazards models for heart failure hospitalization.

|  | Univariable |  |  | Multivariable |  |  |
| --- | --- | --- | --- | --- | --- | --- |
|  | HR | 95% CI | p | HR | 95% CI | p |
| **Age** | 1.02 | 0.99-1.04 | 0.32 |  |  |  |
| **Male Sex** | 0.79 | 0.51-1.22 | 0.30 |  |  |  |
| **Body Mass Index** | 1.02 | 0.97-1.06 | 0.45 |  |  |  |
| **Systolic Blood pressure** | 1.01 | 0.99-1.02 | 0.37 |  |  |  |
| **Hypertension** | 1.19 | 0.64-2.19 | 0.58 |  |  |  |
| **Dyslipidemia** | 1.64 | 1.06-2.54 | 0.03 | 1.38 | 0.84-2.29 | 0.20 |
| **Diabetes** | 1.26 | 0.82-1.94 | 0.30 |  |  |  |
| **Current smoker** | 0.93 | 0.52-1.65 | 0.80 |  |  |  |
| **Antihypertensive drugs** | 2.96 | 0.41-21.23 | 0.28 |  |  |  |
| **RAS inhibitors** | 1.23 | 0.79-1.93 | 0.36 |  |  |  |
| **β blockers** | 1.44 | 0.91-2.26 | 0.12 |  |  |  |
| **MRA** | 0.73 | 0.47-1.16 | 0.18 |  |  |  |
| **Statin** | 1.48 | 0.96-2.29 | 0.08 |  |  |  |
| **Diuretics** | 1.30 | 0.75-2.24 | 0.35 |  |  |  |
| **Red blood cells** | 0.99 | 0.99-1.00 | <0.001 | - | - | - |
| **Hemoglobin** | 0.84 | 0.75-0.93 | 0.001 | 0.89 | 0.80-1.00 | 0.06 |
| **Hematocrit** | 0.94 | 0.90-0.97 | <0.001 | - | - | - |
| **White blood cells** | 0.97 | 0.87-1.09 | 0.60 |  |  |  |
| **Platelets** | 1.00 | 0.97-1.02 | 0.84 |  |  |  |
| **Total protein** | 0.78 | 0.56-1.09 | 0.15 |  |  |  |
| **Albumin** | 0.74 | 0.44-1.25 | 0.26 |  |  |  |
| **AST** | 1.00 | 0.98-1.02 | 0.74 |  |  |  |
| **ALT** | 0.98 | 0.97-1.00 | 0.07 |  |  |  |
| **Sodium** | 0.95 | 0.88-1.03 | 0.21 |  |  |  |
| **Potassium** | 0.97 | 0.91-1.02 | 0.24 |  |  |  |
| **Chloride** | 0.79 | 0.50-1.24 | 0.30 |  |  |  |
| **Blood urea nitrogen** | 1.02 | 1.00-1.04 | 0.04 | 1.00 | 0.97-1.04 | 0.84 |
| **Creatinine** | 1.60 | 1.08-2.37 | 0.02 | 1.28 | 0.66-2.49 | 0.46 |
| **eGFR** | 0.99 | 0.98-1.01 | 0.23 |  |  |  |
| **Blood glucose** | 1.00 | 0.99-1.01 | 0.93 |  |  |  |
| **HbA1c** | 1.05 | 0.79-1.40 | 0.72 |  |  |  |
| **Total cholesterol** | 1.00 | 0.99-1.01 | 0.50 |  |  |  |
| **Triglyceride** | 1.00 | 1.00-1.01 | 0.19 |  |  |  |
| **HDL-C** | 1.02 | 0.99-1.04 | 0.21 |  |  |  |
| **LDL-C** | 1.00 | 0.99-1.01 | 0.84 |  |  |  |
| **C-reactive protein** | 1.11 | 0.92-1.34 | 0.28 |  |  |  |
| **LN(NTproBNP)** | 1.40 | 1.09-1.79 | 0.009 | 1.08 | 0.82-1.43 | 0.58 |
| **Quartile of CVD Risk** |  |  | 0.003 |  |  | 0.27 |
| ***Q2*** | 1.97 | 0.97-4.01 | 0.06 | 2.15 | 0.94-4.89 | 0.07 |
| ***Q3*** | 1.98 | 0.97-4.04 | 0.06 | 1.86 | 0.82-4.25 | 0.14 |
| ***Q4*** | 3.48 | 1.78-6.80 | <0.001 | 1.44 | 0.59-3.52 | 0.43 |

MACE denotes major adverse cardiovascular events; RAS, renin angiotensin system; MRA, mineral corticoid receptor antagonists; AST, aspartate aminotransferase; ALT, alanine aminotransferase; eGFR, estimated glomerular filtration rate; HbA1c, hemoglobin A1c; HDL-c, high-density lipoprotein cholesterol; LDL-c, low-density lipoprotein cholesterol; LN(NT-proBNP), logarithm of N-terminal pro-brain natriuretic peptide; CVD, cardiovascular disease.
MACE

The 10-year HF risk and ASCVD risk estimated by the PREVENT equations showed similar results to those of the CVD risk, except that Q2 of the HF risk was associated with significantly higher risk of stroke (HR 13.5, 95% CI 1.41 - 129.30, p=0.02).

### Incremental Prognostic Value of 10-year CVD Risk

To evaluate the incremental prognostic value of the 10-year CVD risk beyond NT-proBNP, two Cox models for predicting MACE, Model 1 including LN(NT-proBNP) alone and Model 2 incorporating LN(NT-proBNP) and 10-year CVD risk, were constructed. The 10-year CVD risk remained independently associated with MACE after adjustment for LN(NT-proBNP) (HR 1.03 per 1% increase, 95% CI 1.01–1.05, p=0.002). The addition of 10-year CVD risk significantly improved model discrimination (C-index: 0.626 vs. 0.665; ΔC=0.039, p=0.009), and yielded superior model fit (likelihood ratio test p=0.002; AIC: 1138 vs. 1130). These findings indicated that the 10-year CVD risk had a incremental prognostic value for MACE beyond NT-proBNP.

### Comparison with Other Risk Scores

The predictive value of the estimated CVD risk was assessed using time-dependent ROC curves for MACE at 1 year. The AUC of the estimated CVD risk was 0.677 (95%CI: 0.604 - 0.749) (Figure 3). There was no significant difference in AUC between the CVD risk and the HF risk (AUC 0.704, 95%CI: 0.631 - 0.777, p=0.11) or ASCVD risk (AUC 0.657, 95%CI: 0.584 - 0.730, p=0.16) (Figure 3).

**Figure 3.**
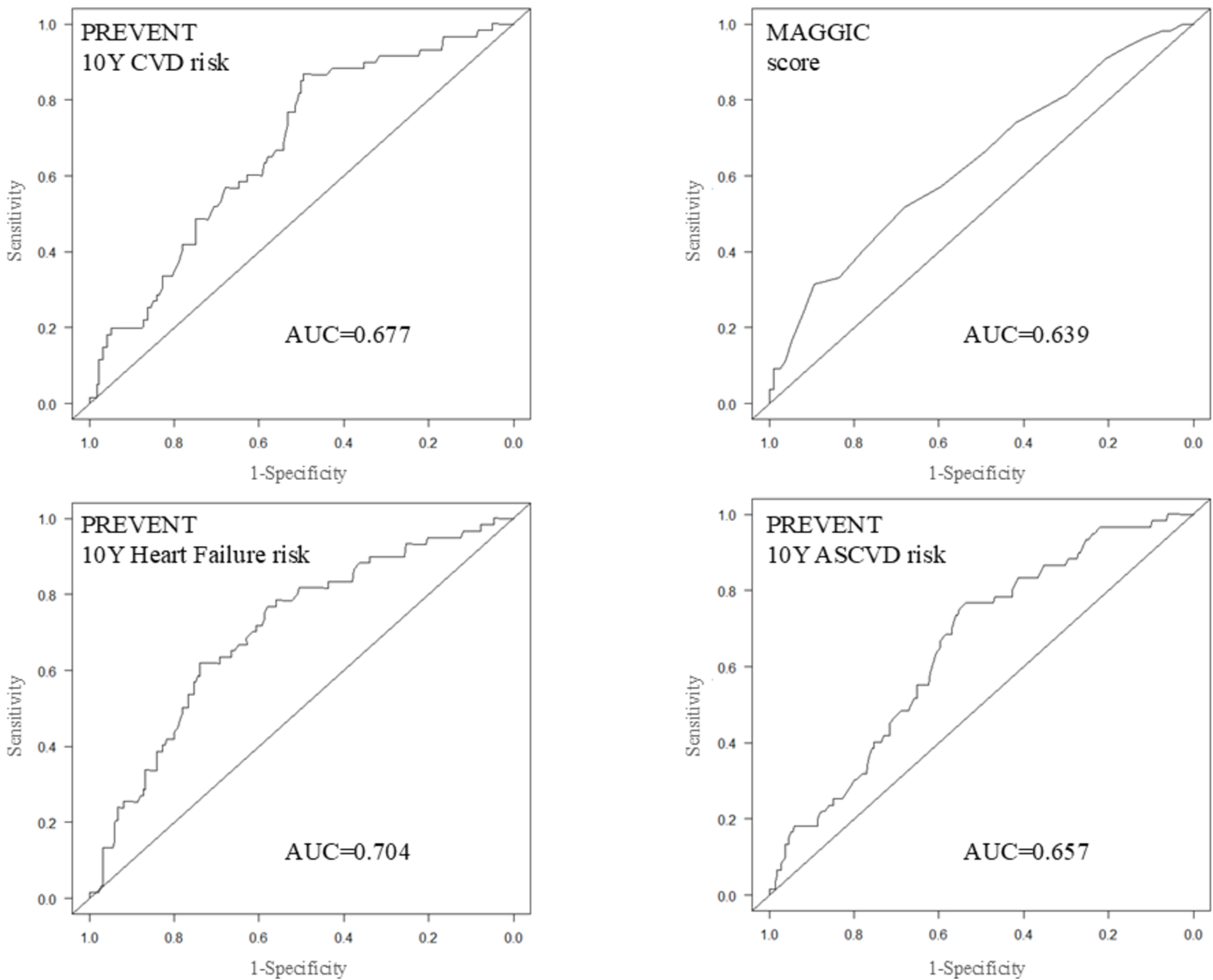
Time-dependent ROC curves for prediction of MACE at 1-year. Time-dependent ROC curves of the 10-year CVD risk (upper left), the MAGGIC score (upper right), HF risk (lower left) and ASCVD risk (lower right) for MACE at 1year after hospitalization. The AUC of the CVD risk (0.677) was not significantly different from that of the MAGGIC (0.639), the HF risk (0.704), or the ASCVD risk (0.657). ROC denotes receiver operating characteristic; MACE, major adverse cardiovascular events; CVD, cardiovascular disease; ASCVD, atherosclerotic cardiovascular disease.

The MAGGIC score could not be calculated in 36 patients, and we compared the AUC of the PREVENT CVD risk score and that of the MAGGIC score in the remaining 242 patients, among whom there were 125 MACE during the follow-up. There was no significant difference in AUC between the estimated CVD risk (AUC 0.676, 95%CI: 0.598 - 0.753) and the MAGGIC score (AUC 0.639, 95%CI: 0.553 - 0.724, p=0.42) (Figure 3).

## Discussion

The present study demonstrated that 10-year CVD risk score estimated by the PREVENT equations in 278 patients hospitalized for decompensated HFpEF was significantly associated with MACE during a median follow-up of 1050 days. The classification by the quartiles of estimated CVD risk was an independent predictor for MACE (p=0.02) in the multivariable Cox proportional hazard model, and the lowest CVD risk quartile (Q1) had a significantly lower incidence of MACE compared with the remaining three quartiles (p< 0.001 by log-rank test). Although there were significant differences in the incidence of all-cause death and HF hospitalization between the four groups, the multivariable Cox analysis did not identify the CVD risk classification as an independent predictor. Although the predictive value of the estimated CVD risk was modest for MACE, it had an incremental value beyond NT-proBNP and was comparable to that of the MAGGIC score. The 10-year ASCVD risk and HF risk estimated by the PREVENT equations showed similar results to those of the CVD risk, and they had similar predictive values for 1-year MACE (AUC 0.657 and 0.704, respectively). These results suggest possible clinical implications of the PREVNT equations for risk stratification in patients with HFpEF.

### The PREVENT 10-year CVD Risk and MACE

The PREVENT equations estimate CVD risk using 12 routinely available cardiovascular risk factors. Whereas significant difference was observed between the four CVD risk groups in 9 of these factors, with the exceptions of systolic blood pressure, total cholesterol, and statin use (Table 1). However, only eGFR and diabetes were identified as independent predictors for MACE in the univariable analysis, and neither was retained in the multivariable analysis. The absence of an independent association between these variables and MACE implies that the predictive value of the estimated CVD risk arises from the aggregation of multiple modest risk contributions, which collectively form a clinically meaningful estimate of overall cardiovascular vulnerability, rather than from any single dominant predictor. Importantly, the PREVENT equations rely solely on general cardiovascular risk factors and do not incorporate any HF-specific variables. The MAGGIC score incorporates HF-relevant parameters such as LVEF, NYHA class, and disease-specific medications to estimate prognosis in HF patients. The AUC of 0.639 for predicting MACE using the MAGGIC score in this study is similar to that reported in previous studies of HFpEF^15–17^. These two scores had a similar predictive value for MACE in this study, suggesting that the burden of general cardiovascular risk factors captured by the PREVENT equations contributes substantially to clinical outcomes even after the clinical onset of HFpEF. The CVD risk in this study cohort was as high as 29.1±9.6 %, ranging up to 22.6 % even in the lowest risk group, indicating that these clinical risk factors influence clinical outcomes even in the very high-risk populations. Management of these clinical factors, paradoxically, may worsen prognosis^16^, as shown in the U-shaped relationship between blood pressure^20^ or HbA1c^16^. It is unclear whether improvement of the clinical factors in the PREVENT equations would lead to better prognosis in patients with HFpEF.

The estimated 10-year CVD risk had an incremental predictive value over NT-proBNP and may offer a practical tool for risk stratification in HFpEF, particularly in settings where HF-specific scoring systems are not readily available. However, its predictive ability remains moderate, and it should not be used as a standalone prognostic tool in this population. It may rather serve as a complementary risk stratification measure alongside HFpEF-specific indices such as the H2FPEF score^18^, HFA-PEFF algorithm^19^, or the PREDICT-HFpEF score^17^, though these scores require HF-specific variables and echocardiography. Further studies are required to compare the prognostic ability of these scores with that of the CVD risk by the PREVENT equations.

The 10-year CVD risk was not identified as an independent predictor for the individual components of MACE. The number of events for each component outcome was smaller than that for MACE, which may have reduced statistical power in multivariable analyses. The PREVENT equations primarily reflect long-term (10- and 30-years) cardiovascular risk rather than short-term hemodynamic instability, which is a major driver of HF hospitalization. Anyway, the PREVENT equations are better suited to capture a global vulnerability to MACE rather than predicting its individual component with high specificity.

The primary objective of this study was to predict overall prognosis in HFpEF as a composite endpoint. Non-cardiovascular death and stroke substantially contribute to outcomes in HFpEF^27–31^, and we hypothesized that the risk for total CVD would be related to the total clinical outcome and used it in the present study. The comparison of time-dependent ROC curves indicated that 10-year risk for incident HF and ASCVD had similar predictive value for MACE at 1 year. However, we did not analyze whether these risk factors could predict all-cause death or HF hospitalization better than the CVD risk. ASCVD risk and HF risk reflect distinct pathophysiological domains, and their potential prognostic value for specific outcomes cannot be excluded. Further studies are warranted to clarify their role in predicting individual clinical endpoints in HFpEF.

### Applicability of the PREVENT Equations

One of the critical issues of this study is the applicability of the PREVENT equations to the study cohort. The final study population consisted of 278 patients, representing only 22.4% of the initial 1238 patients, and the small sample size may have introduced selection bias into this study. The PREVENT equations are applicable to individuals aged 30 to 79 years. The median age of the registered patients was 83 years (IQR 77, 87), and only 433 out of 1238 patients (35%) were within this range. The age distribution of HF patients varies considerably across regions, and Japanese patients tend to be among the oldest globally. The mean age in the nationwide registries such as the Japanese Registry Of Acute Decompensated Heart Failure (JROADHF) was 78.0±12.5 years^32^, and the Japanese Heart Failure Syndrome With Preserved Ejection Fraction (JASPER) Registry had a median age of 80 years^33^, both of which are compatible with that of the PURSUIT-HFpEF registry. The age of Japanese HF patients tends to be higher than those of not only that of Western countries^4,27,34^ but also other Asian countries^35^, reflecting Japan’s aging population.

The next major cause of patient exclusion was variables outside the acceptable range for CVD risk calculation, which excluded 155 (35.8%) out of 433 patients with an eligible age. The out-of-rang variables included low total cholesterol in 42 patients, low BMI in 58 patients, and low eGFR in 36 patients, which also reflect the characteristics of Japanese HFpEF patients. Japanese HFpEF patients have relatively low BMI, and the obesity rates are characteristically low compared with Western populations^33,36^. Low total cholesterol may be related to lean body composition and older age. Although the prevalence of CKD in Japanese HFpEF patients is reported to be comparable to that in other countries^37^, it has been increasing in recent years, probably owing to population aging^3^. As expected, the patients excluded for the analysis had lower BMI, worse nutritional status (lower albumin and total protein), more severe renal impairment, more pronounced anemia, lower lipid levels, and higher NT-proBNP levels, suggesting a generally sicker phenotype (Supplement Table S1). The applicability of the PREVENT equation may be limited in HF patients with such unfavorable clinical profile, especially in East Asian population.

### Study Limitations

The present study has several other study limitations other than the applicability of risk estimation. This is a retrospective study and is subject to several types of bias including selection bias and confounding. The PREVENT equations are designed to calculate 10-and 30-year risk, and the follow-up period of this study may have been too short to evaluate the full predictive value of the estimated CVD risk. Urine albumin-to-creatinine ratio, which was not recorded in the registry database, was not used for the risk calculation, and HbA1c was not available in 18% of patients, though they were optional for the calculation. The PREVENT equations are race-free and their ability to predict incidental CVD, ASCVD, and HF has been confirmed in Asian population. However, there are substantial regional and international differences in the characteristics of HFpEF patients, reflecting variations in demographics, comorbidity profiles, and contributing risk factors, and it is uncertain whether the present results are generalizable to other ethnic populations, particularly Western populations. Despite these limitations, the present study demonstrated the significant influence of CVD-related risk factors on clinical outcomes in patients hospitalized with HFpEF and suggested a possible role for the PREVENT equations in stratifying the high-risk populations within this patient group.

## Data Availability

The data that support the findings of this study are available from the corresponding author upon reasonable request.

